# The association between loneliness during the COVID-19 pandemic and psychological distress

**DOI:** 10.1101/2021.04.08.21255118

**Authors:** Yusuke Konno, Masako Nagata, Ayako Hino, Seiichiro Tateishi, Mayumi Tsuji, Akira Ogami, Reiji Yoshimura, Shinya Matsuda, Yoshihisa Fujino, for the CORoNaWork Project

## Abstract

The purpose of this study was to examine the association between loneliness and psychological distress during the COVID-19 pandemic in Japan. We conducted a cross-sectional, online study from 22 to 26 December 2020. A total of 27,036 participants, all employed at the time of the survey, were included in the analysis. Participants were asked if they felt loneliness in a single-item question. The Kessler 6 (K6) was used to assess psychological distress defined as K6 scores of 5 or higher, and 13 or higher. The odds ratios (ORs) of psychological distress associated with loneliness were estimated using a multilevel logistic model nested in the prefecture of residence, with adjustment for age, sex, marital status, equivalent income, educational level, smoking, alcohol consumption, job type, number of workplace employees, and cumulative incidence rate of COVID-19 in the prefecture. Communication with friends, acquaintances, and family was strongly associated with psychological distress, so we adjusted for these factors and eating meals alone. Results showed a significant association between loneliness and psychological distress (OR = 36.62, 95%CI = 32.95-40.69). Lack of friends to talk to, lack of acquaintances to ask for help, and lack of people to communicate with through social networking sites were all strongly associated with psychological distress, as were family time and solitary eating. Even after adjusting for these factors, loneliness was still strongly associated with psychological distress (OR = 29.36, 95%CI = 26.44-32.98). The association between loneliness during the COVID-19 pandemic and psychological distress indicates the need for intervention.

## 1. Introduction

The outbreak of the new coronavirus infection (COVID-19) caused by SARS-CoV-2 was first reported in Wuhan, China, in December 2019, and it has since spread around the world. The World Health Organization (WHO) declared a public health emergency on 30 January 2020, and on 11 March 2020, declared it a pandemic. In Japan, the first case was reported on 16 January 2020, and the disease was designated as a designated infectious disease on 1 February 2020. As of March 2021, the disease continues to spread around the world. Japan is no exception, and since the first case was reported, the outbreak has spread, with a cumulative total of 324,846 confirmed cases in the year from 16 January 2020 to 16 January 2021.^1^ In addition to its infectiousness, COVID-19 was found to be highly contagious, with high severity of illness and a high mortality rate in severely ill patients, leading to measures aimed at preventing COVID-19 in many countries around the world.^2^

In its “Coronavirus disease (COVID-19) advice for the public” to combat COVID-19, WHO advised to: “Maintain at least a 1-meter distance between yourself and others to reduce your risk of infection when they cough, sneeze or speak. Maintain an even greater distance between yourself and others when indoors.” It also recommends avoiding the 3Cs, i.e. spaces that are closed, crowded, or involve close contact, using the phrase: “Avoid the 3Cs”.^3^

These COVID-19 measures were also applied in Japan, resulting in a transformation of lifestyles and making people more isolated. The Ministry of Health, Labour and Welfare (MHLW) has announced a “new lifestyle” for the new coronavirus, recommending that people avoid contact with other people as much as possible, and if they do make contact, it should be for short periods of time; they should avoid talking, and maintain as much distance from others as possible. People are advised to refrain from going out unnecessarily and to shop online as much as possible.^4^ Living the kind of life recommended for COVID-19 prevention limits communication and makes it easier to socially withdraw.

The work environment is no exception with respect to COVID-19 measures resulting in greater isolation. “Examples of ‘new lifestyle’ practices in anticipation of the new coronavirus” recommend telework and online meetings, rota working, and staggered commuting.^4^ As a result, employees work alone at home, and meetings are held on the web. They no longer have dinners with their workmates, but rather eat and drink alone. No more crowded commuting, no more being with others in enclosed spaces such as offices or conference rooms, no more conversations without wearing a mask when eating or drinking. In parallel, opportunities for both public and private communication have decreased, and people’s lifestyle has become more solitary than before the COVID-19 pandemic.

It has been reported that during the COVID-19 pandemic, people lead more isolated lives and that feelings of psychological distress have increased due to COVID-19 prevention measures.^7,8^ Psychological distress causes physical and mental health problems. Physically, psychological distress can trigger the onset and exacerbation of cardiovascular diseases such as hypertension, arrhythmia, ischemic heart disease and heart failure, and chronic stress can lead to overeating and lack of exercise, which in turn leads to obesity, placing an additional burden on the cardiovascular system.^9^ Furthermore, excessive psychological distress is a risk factor for mental disorders including depression.^10^

The longer the COVID-19 pandemic continues, the more psychological distress is expected to increase, and the more physical and mental disorders caused by psychological distress are expected to increase; therefore, we believe that psychological distress alleviation measures are necessary. The causes of psychological distress resulting from the COVID-19 pandemic are thought to be diverse, but if one of them is isolated living and the sense of loneliness that arises from it, then countermeasures are necessary. The purpose of this study is to examine the association between loneliness and psychological distress under COVID-19 conditions, and to help improve health.

## 2. Methods

### 2.1. Study design and participants

This cross-sectional study was conducted on the Internet from 22 to 26 December 2020. The details of the protocol have already been reported.^11^ In brief, data were collected from people who were in employment at the time of the survey, selected by prefecture, occupation, and gender. A total of 33,302 people participated in the survey. After excluding clearly untruthful responses, data from 27,036 participants were included in the analysis.

### 2.2. Assessment of loneliness

In this survey, one question was used to determine whether the participants felt loneliness or not. To the question: “During the last 30 days, how frequently have you felt loneliness?”, the subjects answered by selecting one option from: never, a little, sometimes, usually, always. If the subject answered always, usually, or sometimes, loneliness was considered to be present.

### 2.3. Assessment of psychological distress

Kessler 6 (K6) was used to assess psychological distress.^12^ The validity of the Japanese version of K6 has been confirmed.^5,6^ In this study, the cutoff for mild psychological distress was a K6 score of 5 or higher, and for severe psychological distress, a score of 13 or higher.

### 2.4. Other covariates

The following survey items were considered confounding factors: age, sex, marital status, equivalent income, educational level, smoking, alcohol consumption, job type, number of employees at the workplace and cumulative incidence rate of COVID-19 in the prefecture. The survey also asked questions such as: “Do you have friends or neighbors with whom you can easily engage in small talk or daily conversation? “; “Do you have someone you can ask for help?” and “Do you have a partner with whom you can communicate closely using SNSs?” The participants responded “Yes” or “No” to these questions. For the question: “Time spent with family having a meal or at home”, participants answered: more than 2 hours, more than 1 hour, more than 30 minutes, less than 30 minutes, almost never. For the question: “How often do you eat all meals of the day alone?” they answered: 6–7 days a week, 4–5 days a week, 2–3 days a week, less than 1 day a week, hardly ever.

In addition, the cumulative incidence of COVID-19 from the time of the survey to one month before in the prefecture of residence was used as a community-level variable. Information was collected from the websites of public institutions.

### 2.5. Statistical analysis

Odds ratios (ORs) for loneliness and psychological distress were estimated with a logistic model. Psychological distress was defined as mild psychological distress with a K6 score of 5 or higher, and severe psychological distress with a K6 score of 13 or higher. In the multivariate model, we adjusted for age, sex, marital status, equivalent income, educational level, smoking, alcohol consumption, job type, number of employees in the workplace, and cumulative incidence rate of COVID-19 in the prefecture. In another model, we added having friends or neighbors with whom to easily make small talk or have daily conversations, having someone who can be asked for a little help, and having a close friend to communicate with on social networking sites. The rate of COVID-19 incidence by prefecture was used as a prefecture-level variable.

A p-value of less than 0.05 was considered statistically significant. Stata (Stata Statistical Software: Release 16. College Station, TX: StataCorp LLC.) was used for analysis.

## 3. Results

The characteristics of the database are shown in table 1. Of the 27,036 participants, 2,750 (10%) felt loneliness. Of those who reported feeling loneliness, 93.6% had mild psychological distress and 58.5% had severe psychological distress. In contrast, only 33.9% of the group who did not feel loneliness had mild psychological distress and 3.5% had severe psychological distress. The percentage of participants who answered “yes” to the questions: ”Do you have friends or neighbors with whom you can easily engage in small talk or daily conversation?”, ”Do you have someone you can ask for help?”, and ”Do you have a partner with whom you can communicate closely using SNSs?” was lower in the loneliness group in all cases. Those who spent less time with their family during meals and gatherings were more likely to feel loneliness, while those who ate all their meals alone were more likely to feel loneliness. The percentage of participants who felt loneliness was higher among women, the unmarried, and those on low incomes. No differences emerged in relation to job type or number of employees in the workplace.

Table 2 shows the odds ratios (ORs) of loneliness and severe psychological distress estimated by the logistic model. In the age-adjusted model, there was a significant association between loneliness and psychological distress (OR = 37.74, 95%CI = 34.04-41.85). This result was also found in the multivariate analysis (OR = 36.62, 95%CI = 32.95-40.69). Lack of friends to talk to, lack of acquaintances to ask for favors, and lack of people to communicate with through social networking sites were all strongly associated with psychological distress. Family time and solitary eating were both associated with psychological distress. Even after adjusting for these factors, loneliness was still strongly associated with psychological distress (OR = 29.36, 95%CI = 26.44-32.98).

## 4. Discussion

In this study, 10% of all participants felt loneliness, and this loneliness was strongly associated with psychological distress. Other studies have reported that people’s psychological distress increased during the COVID-19 pandemic.^13^ Communication with friends, acquaintances, and family was also strongly associated with psychological distress; we therefore adjusted for spending time with friends, acquaintances, and family, but loneliness was still strongly associated with psychological distress. Loneliness is generally defined as the discrepancy between a person’s desired social relationships and their actual social relationships.^14^ And loneliness is considered to be different from social isolation, although there is overlap between the two.^14^ In this study, loneliness was assessed by the subjective question: “Have you ever felt loneliness?” By contrast, communication with friends, acquaintances, and family, which was used as an adjustment factor, was assessed using objective questions, as was social isolation. The results of this study showed that loneliness and social isolation overlap, but only partially, and the non-overlapping parts are considered to be subjective experiences. We believe that this subjective experience is associated with psychological distress and is a factor that should be addressed.

One of the reasons for the loneliness highlighted here is considered to be the particular lifestyle that has arisen due to the COVID-19 pandemic. This lifestyle requires a different way of living, complying with “Avoid the 3Cs”^15,16^ to prevent infection, but as a result, communication has decreased. This has led to a sense of loneliness, which in turn is thought to be linked to psychological distress. As of February 2021, there is no prospect of COVID-19 eradication, and people will need to continue their new lifestyles to prevent infection. Therefore, the incidence of psychological distress is expected to remain high for some time. As severe psychological distress conditions can be physically and mentally disabling, interventions for loneliness are needed if psychological distress under COVID-19 pandemic conditions persists.

As an intervention for loneliness, the usefulness of online communication such as social media has been advocated.^17^ Online communication tools, such as social networking sites, can be a valuable countermeasure against loneliness during the pandemic, and are also useful in terms of infection prevention.^18^ In a randomized controlled trial conducted during the COVID-19 pandemic, an empathy-focused program of telephone calls significantly improved loneliness.^19^ Thus, various types of online communication, such as communication via the web and telephone calls, and using social media, may be useful as interventions for loneliness during the COVID-19 pandemic.

However, it has also been pointed out that continual use of social media and other forms of online communication in situations of loneliness may be harmful. One risk is increased psychological distress caused by excessive media consumption. Being inundated with inaccurate information and excessive media consumption during the COVID-19 pandemic may lead to deteriorating mental health.^20^ WHO has cautioned that with the current growing use of social media and the internet, not only useful but also inaccurate or harmful information about COVID-19 can spread quickly, leading to confusion, health problems and mistrust of health institutions.^21–23^

Another risk is the issue of addiction. Loneliness is a risk factor for substance or behavioral addictions, including the Internet.^24,25^ In addition, addiction to the Internet increases one’s loneliness, and the increased loneliness worsens the addiction, thus creating a vicious cycle.^26^

Thus, there is a risk that online communication, even when used with the intention to reduce psychological distress, may conversely exacerbate or cause such distress, or lead to addictive behaviors. Online communication is useful as a countermeasure to loneliness, but in some cases, it may have harmful consequences, and therefore it should be used with caution.

### 4.1. Limitations of the study

There are several limitations to this study. First, because it was conducted online using the Internet, the generalizability of the results is uncertain, but we attempted to minimize participant bias by sampling by occupation, region, and prefecture based on the incidence of infection. Second, the accuracy of the reported incidence of loneliness may be questioned because loneliness was evaluated only by the question: “Have you ever felt loneliness?” In studies that evaluated accuracy, the binary evaluation of living alone had the lowest accuracy.^26^ However, we feel that the question asked in this study is appropriate, as it briefly asks about participants’ subjective experiences.

Third, because this is a cross-sectional study, the temporal relationship between loneliness and psychological distress is unknown.

## 5. Conclusion

We found that 10% of the participants felt loneliness living under conditions of the COVID-19 pandemic. Loneliness was associated with psychological distress, which we believe requires intervention. Online communication is considered to be an effective intervention in loneliness, but at the same time, it is important to take into account risks, such as addiction.

## Data Availability

N/A

## Acknowledgments

This study was supported by a research fund from the University of Occupational and Environmental Health, Japan; General Incorporated Foundation (Anshin Zaidan); The Development of Educational Materials on Mental Health Measures for Managers at Small-sized Enterprises; Health, Labour and Welfare Sciences Research Grants; Comprehensive Research for Women’s Healthcare (H30-josei-ippan-002); Research for the Establishment of an Occupational Health System in Times of Disaster (H30-roudou-ippan-007), and scholarship donations from Chugai Pharmaceutical Co., Ltd.

Present members of the Collaborative Online Research on the Novel-coronavirus and Work (CORoNaWork) Project are: Dr. Yoshihisa Fujino (current chairperson), Dr. Akira Ogami, Dr. Arisa Harada, Dr. Ayako Hino, Dr. Chimed-Ochir Odgerel, Dr. Hajime Ando, Dr. Hisashi Eguchi, Dr. Kazunori Ikegami, Dr. Keiji Muramatsu, Dr. Koji Mori, Dr. Kyoko Kitagawa, Dr. Masako Nagata, Dr. Mayumi Tsuji, Dr. Rie Tanaka, Dr. Ryutaro Matsugaki, Dr. Seiishiro Tateishi, Dr. Shinya Matsuda, Dr. Tomohiro Ishimaru, Dr. Tomohisa Nagata, Dr. Yosuke Mafune, and Ms. Ning Liu, in alphabetical order. All of the members are affiliated with the University of Occupational and Environmental Health, Japan.

